# Effects of long-chain omega-3 polyunsaturated fatty acids on reducing anxiety and/or depression in adults; A systematic review and meta-analysis of randomised controlled trials

**DOI:** 10.1101/2022.10.17.22281092

**Authors:** Christos F. Kelaiditis, E. Leigh Gibson, Simon C. Dyall

## Abstract

First-line treatment for anxiety and depressive disorders comprises pharmacotherapy and psychotherapy; options not safe, effective, or suitable for all. Mounting evidence suggests that the omega-3 polyunsaturated fatty acids (PUFAs) eicosapentaenoic (EPA), docosahexaenoic (DHA) and docosapentaenoic (DPAn-3) acids are promising therapeutic options. However, meta-analyses of randomised controlled trials (RCTs) have produced inconsistent findings. This review assesses for the first time the efficacy of omega-3 PUFAs against the severity of anxiety and depression symptoms, measured by validated scales, with specific consideration of methodological issues encountered in this area. PubMed, CINAHL, PsycINFO, Cochrane Library and Web of Science were searched for eligible RCTs administering omega-3 PUFAs against anxiety and/or depression. This study adopts the PRISMA guidelines. Ten RCTs comprising 1509 participants were included in the quantitative synthesis. EPA-enriched interventions at ≥60% of total EPA+DHA were associated with significant reduction in depression severity, compared to placebo (SMD: −0.32; 95% CI: − 0.59, −0.06; p=0.02); however, EPA doses of ≥2000 mg/day were not (SMD: − 0.11; 95% CI: −0.43, 0.20; p=0.48). Only 10 RCTs fulfilled the eligibility criteria, and there were some concerns regarding bias and population heterogeneity, highlighting the lack of high-quality RCTs in this area. Overall, these results support previous observations where EPA at proportions ≥60% of total EPA+DHA, up to 2000 mg, reduces depression scores. However, more trials are needed which specifically consider the unique nature of this type of research to elucidate the therapeutic potential of EPA, DHA and DPAn-3.

## Introduction

Anxiety and depressive disorders were the most debilitating mental disorders in 2019 [1], with over 300 million cases of anxiety, and 280 million cases of depression worldwide [2]. By 2020, the cases of anxiety and major depressive disorder (MDD) increased by 25.6% and 27.6% respectively due to the COVID-19 pandemic [3]. First-line treatment for anxiety and depressive disorders comprises pharmacotherapy and psychotherapy [4], options which may not be safe, effective, or suitable for all [5]. In contrast, diet might be a key modifiable environmental factor which modulates the onset and progression of these affective disorders [6].

The omega-3 polyunsaturated fatty acids (PUFAs) eicosapentaenoic acid (EPA) and docosahexaenoic acid (DHA) are key dietary components investigated extensively in relation to psychiatric outcomes, owing to the therapeutic potential of their ever-expanding repertoire of potent bioactive derivatives [7]. Despite the different bioactivity between EPA and DHA, there is a robust biological basis supporting their anxiolytic and antidepressant potential, comprising modulation of neuronal membrane properties, receptor expression and neurotransmission, antioxidant, anti-inflammatory and pro-resolving activity, and promotion of neuroplasticity and neuroprotection [8]. These properties may modulate brain homeostasis, which in turn is linked to mood [9], making it imperative to understand the role of omega-3 PUFAs in mental health.

Docosapentaenoic acid, in its n-3 form (DPAn-3), is also gaining recognition as a key bioactive molecule implicated in neuroinflammation [8] and mental health outcomes [10]. In animal models, supplementation with DPAn-3 improves depressive symptoms [11], while in human prospective studies, increased dietary DPAn-3 intake is associated with reduced risk of MDD [12]. However, this PUFA is often neglected by the therapeutic research and typically excluded from systematic reviews and meta-analyses. Preclinical and epidemiological studies consistently associate anxious and depressive states with poor omega-3 PUFA status [13-15]. Correcting the latter by supplementation improves mood profiles [16]. However, meta-analyses of RCTs administering omega-3 PUFAs have produced mixed results. Some report little or no effect in preventing depressive or anxiety symptoms [17], while others report clear anxiolytic and antidepressant effects, accentuated in clinical populations, at higher doses of EPA; but not DHA [18-20]. These discrepancies can be attributed to methodological limitations encountered in omega-3 PUFA research, comprising heterogeneity between studies and potential bioactivity of placebo preparations [21] and the influence of background diet and interindividual responsiveness to supplementation and genotype variation [22].

A particularly important limitation of research in this area is the lack of discrimination between the different omega-3 PUFAs and appreciation of their unique and individual biological activities [8]. EPA and DHA may counter-modulate their respective physiological effects on membranes and compete with each other for incorporation into membrane phospholipids, with further implications for membrane architecture, intracellular signalling, and biosynthesis of bioactive mediators, suggesting that simultaneous administration of these omega-3 PUFAs may confound outcomes [23]. Aberrant inflammatory processes have been implicated as potential pathophysiological mechanisms and treatment targets in mood disorders [24], and although omega-3 PUFA-derived oxylipins display potent anti-inflammatory and pro-resolving properties that may underlie their therapeutic effects, there are differences between the activities of the mediators formed from each of the respective omega-3 PUFAs [7].

Early meta-analyses of RCTs administering omega-3 PUFA regimes comprising ≥50% DHA, with EPA, suggested that DHA-enriched regimes are not effective against depression, leading to the hypothesis that DHA does not improve depressive symptoms [25, 26], and may also inhibit the antidepressant bioactivity of EPA by slowing down its conversion rate to downstream bioactive metabolites [27]. In comparison, meta-analysis of RCTs administering EPA-enriched regimes demonstrated that EPA at proportions ≥60% of total EPA+DHA, in the dose range 200-2200 mg are effective against primary depression [20]. However, more recent meta-analyses suggest omega-3 PUFA doses of ≥2000 mg confer anxiolytic effects at proportions of ≤60% EPA; but not at EPA-enriched proportions of ≥60% EPA [19]. The present systematic review and meta-analysis investigates this discrepancy in the anxiolytic/antidepressant potential of EPA to explore whether there is an optimal therapeutic dose and ratio of EPA:DHA.

This study assessed the efficacy of omega-3 PUFAs supplementation in improving anxiety and depression outcomes against placebo, by identifying and synthesising evidence from high-quality RCTs administering EPA and/or DHA and/or DPAn-3, either as monotherapies or adjuvants, in adults exhibiting various levels of anxiety and/or depression. It builds on previous findings [19, 20], focusing on RCTs administering EPA-enriched fish-oil regimes for at least eight weeks, to determine whether EPA at high doses of ≥2000 mg/day, or proportions ≥60% of total EPA+DHA, reduces the severity of anxiety and/or depression symptoms in adults. Specifically, this systematic review and meta-analysis sought to answer the following questions: (a) Does EPA at proportions ≥ 60% of total EPA+DHA reduce the severity of anxiety and depression symptoms in adults? (b) Does EPA at doses ≥2000 mg/day reduce the severity of anxiety and depression symptoms in adults? (c) What is the evidence for the therapeutic effects of DPAn-3?

To minimise confounding caused by the presence of bioactive components in placebo preparations, RCTs were only considered eligible when administering placebo regimes neutral in relation to the outcome i.e., free of bioactive lipids e.g., olive oil [21]. To minimise confounding further, studies were excluded wherever they comprised populations presenting lipid-profile altering conditions e.g., type 2 diabetes mellitus, polycystic ovary syndrome, liver disease and hepatitis, which may confound red blood cell PUFA levels [28], which is the biomarker of omega-3 PUFA status typically utilised by RCTs administering omega-3 PUFA preparations.

## Methods

### Reporting and protocol

This systematic review and meta-analysis adopted the Preferred Reporting Items for Systematic Reviews and Meta-analyses (PRISMA) reporting guidelines [29] and is registered with the International Prospective Register of Systematic Reviews (PROSPERO, ID: CRD42020194402). The PICO elements comprise:

#### Population

Adults ≥18 years of age for whom there is a validated measure of anxiety and/or depression. Excluded were pregnant women, patients with lipid-profile altering diseases/conditions e.g., type 2 diabetes mellitus, polycystic ovary syndrome, liver disease, hepatitis, and severely unwell patients e.g., participants with severe injury, cancer diagnosis, dementia.

#### Intervention

Any supplemental form of long chain omega-3 PUFA comprising EPA with/without DHA and/or DPAn-3 with EPA at high doses of ≥2000 mg/day, or proportions ≥60% of total EPA+DHA administered for ≥8 weeks.

#### Comparator

Placebo preparations reflecting typical dietary intakes of fatty acids of respective countries in which RCTs were conducted, or placebo preparations comprising inert, non-bioactive constituents in relation to the outcome. Excluded were placebo preparations comprising bioactive lipids e.g., olive oil.

#### Outcomes

Baseline to endpoint standardised mean differences (SMD) and/or endpoint scores of severity of anxiety and/or depression measured by validated tools. The abbreviation ‘SMD’ is used interchangeably with ‘effect size’; referred to as Cohen’s d in the literature [30]; SMD values of 0.2-0.5 refer to small, 0.5-0.8 to medium, and >0.8 to large effect sizes respectively [31].

### Search strategy

The literature was searched systematically for eligible studies on PubMed (MEDLINE), CINAHL, PsycINFO, Cochrane Library and Clarivate Web of Science up to 25^th^ July 2022, focusing on peer-reviewed articles written in English, without publication-date restrictions. All searches were performed electronically, and the search strategy syntaxes for the respective databases are reported at the international prospective register of systematic reviews (https://www.crd.york.ac.uk/PROSPEROFILES/194402_STRATEGY_20201126.pdf). The grey literature was searched in reference lists of previously published systematic reviews and meta-analyses in the field to identify manually additional eligible RCTs [15-17, 19, 20, 32-43], the British Library EthOS database, the DART-Europe E-theses Portal, Google Scholar, the World Health Organization’s International Clinical Trials Registry Platform and ClinicalTrials.gov.

### Eligibility criteria

Included RCTs assessed anxiety and/or depression as either primary or secondary outcomes using validated tools i.e., Hamilton Depression Rating Scale (HDRS), Beck Depression Inventory (BDI), Beck Anxiety Inventory (BAI), Center for Epidemiologic Studies Depression Scale (CES-D), Inventory of Depressive Symptomatology Self Report (IDS-SR), Montgomery-Asberg Depression Rating Scale (MADRS), Hopkins Symptom Checklist for Depression (HSCL-D) and Geriatric Depression Scale (GDS). Eligible studies comprised participants, with or without current- or past diagnoses of anxiety- or depressive disorders, supplemented with EPA with/without DHA and/or DPAn-3 preparations as monotherapies or adjuvants. Studies in pregnant women, patients with lipid altering conditions (e.g., type 2 diabetes mellitus, polycystic ovary syndrome, liver disease, hepatitis) and severely ill patients (e.g., participants with severe injury, cancer diagnosis, dementia) were excluded. Studies were also excluded wherever they administered placebo regimes rich in bioactive components (e.g., olive oil and omega-3 PUFAs), had no placebo arm, were duplicate publications, were RCTs following mixed study designs (e.g., a double-blind phase followed by an open-label phase), or were RCTs with several methodological discrepancies and low completion rates. Non-intervention studies, non-RCT interventions and studies investigating several risk factors and/or interventions other than omega-3 PUFAs were also excluded.

Two reviewers (CK and SCD) conducted study eligibility checks independently. After an initial removal of duplicate records, identification of eligible studies was performed by title and abstract review, followed by removal of ineligible studies (with reasons) and full-text reading of the eligible remainders. Wherever disagreements occurred, they were resolved through discussion or mediation by a third reviewer (LG).

### Risk of bias

Two reviewers (CK and SCD) independently assessed risk of bias using the Revised Cochrane risk-of-bias tool for randomized trials (RoB2) [44] which comprises five domains i. randomization process, ii. deviations from the intended interventions (effect of assignment to intervention), iii. missing outcome data, iv. measurement of the outcome and v. selection of the reported result. These domains informed the overall risk of bias judgement with three possible outcomes i.e., low risk, some concerns, and high risk of bias. Wherever disagreements occurred, they were resolved through discussion or mediation by a third reviewer (LG).

### Data extraction

Data was extracted for post-intervention mean scores (adjusted and unadjusted), change-from-baseline scores and their respective standard deviations (SD) for validated outcome measures from eligible studies focusing on intention-to-treat data. Based on a pragmatic approach, authors of studies with missing data were not contacted and missing values were calculated with the Cochrane Review Manager 5.4 (RevMan) tool and the WebPlotDigitizer.

### Heterogeneity and publication bias

Between-trial heterogeneity was assessed with *I*^*2*^ [45] and publication bias by visual examination of funnel plots [46].

### Data synthesis and subgroup analysis

Random-effects meta-analysis was performed using RevMan 5.4 due to heterogeneity in the eligible studies [47], and the primary comparison was the efficacy of supplementation with EPA at high doses of ≥2000 mg/day, or proportions ≥60% of total EPA+DHA on validated measures of anxiety and/or depression vs. placebo (no studies were identified for interventions with DPAn-3). The respective results were standardised to compare the effects of the interventions on anxiety and/or depression. The estimate of the intervention effect for each study relative to the between-participant variability in outcome measurements in the respective studies was reported as SMDs. The pooled summary effect and 95% confidence intervals (CI) were calculated on a conservative random-effects basis to account for the use of different outcome measures by the respective RCTs. Meta-analysis was only performed where there was data for at least two studies. Wherever studies comprised more than one active intervention arms, only the data corresponding to the highest dose was included in the meta-analyses, as it would not be meaningful to meta-analyse data on low doses. Subgroup analyses pooled the effect sizes of studies depending on their respective reporting styles i.e., studies reporting their findings as estimated means at endpoint vs. mean changes from baseline.

## Results

The combined search strategy revealed 791 articles, Fig 1. Of those, 158 duplicates were removed, 571 articles were excluded after title and abstract screening, 62 articles were assessed by full-text reading and 52 were excluded with reasons. Ten studies were included in the quantitative analysis with a mean duration of 11 weeks and a total of 1509 participants (68% female) of a mean age of 46.9 years [48-57]. The effects of DPAn-3 on measures of anxiety or depression were not assessed by any identified trial.

**Fig. 1.**
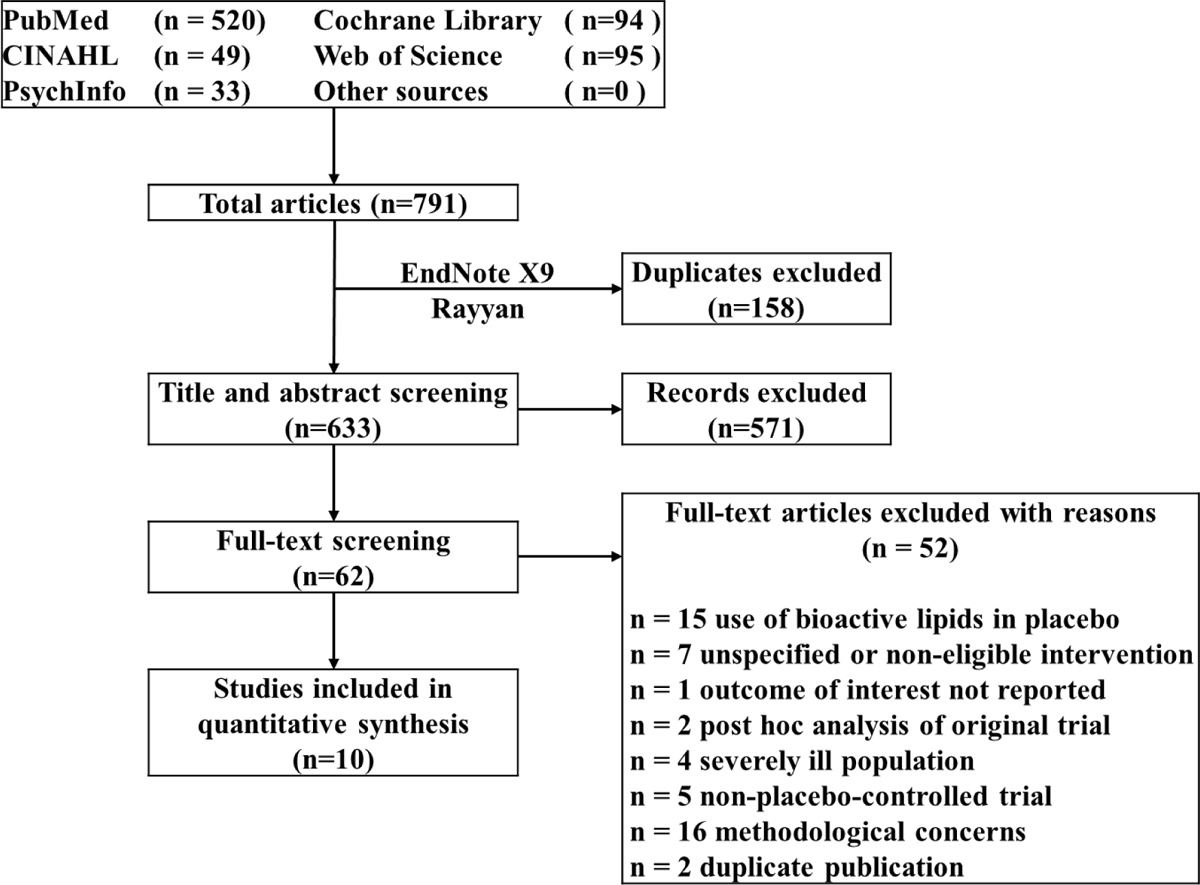
PRISMA flow diagram outlining study selection and exclusion.

### Study characteristics

Table 1 shows the included studies’ characteristics. The severity of anxious and/or depressive symptoms following EPA with/without DHA supplementation were assessed by several validated scales comprising HDRS, BDI, BAI, CES-D, IDS, MADRS, HSCL and GDS. Participants were generally healthy (non-clinical) adults in three RCTs [49-51]. The remainder of the RCTs comprised adults with various degrees of severity of depression and/or anxiety [48, 52-57]. Five RCTs compared EPA-enriched, mixed preparations of EPA/DHA vs. placebo [49, 50, 52, 53, 57]. Of the remaining RCTs, one compared low vs. high dose of EPA vs. placebo [48], one compared equal doses of EPA vs. DHA vs. placebo [55], one compared high vs. low EPA-enriched, mixed preparations of EPA and DHA vs. placebo [51] and two compared EPA-enriched vs. DHA-enriched mixed preparations of EPA and DHA vs. placebo [54, 56]. In the majority of the RCTs, placebo comprised oils of vegetable origin e.g., soybean, sunflower and coconut oils [49, 52-56]. Two of the RCTs administered paraffin oil as a placebo [48, 57]. Of note, two RCTs administered a mixed-fat placebo regime comprising palm, soy, canola, coco butter and olive oil, to approximate the saturated:monounsaturated:polyunsaturated fat ratio consumed by adults in the U.S.A. [50, 51]. Due to the mixed-fat nature of this placebo the reviewers considered the dose of olive oil provided to be insufficient to confound the analysis and consequently the two trials were considered eligible for inclusion.

**Table 1.** Eligible Studies Characteristics.

| **Study** | **Duration (weeks)** | **Mean Age (years)** | **Female (%)** | **Population** | **Intervention** | **Comparator** | **Outcome** |
| --- | --- | --- | --- | --- | --- | --- | --- |
| Frangou 2006 | 12 | 47.0 | 76.0 | Adults with bipolar depression | 1000 mg EPA OR 2000 mg EPA | paraffin oil | HDRS |
| Ginty 2017 | 18 | 43.1 | 50.7 | Healthy adults | 1000 mg EPA AND 400 mg DHA | soybean oil | BDI |
| Kiecolt-Glaser 2011 | 12 | 23.6 | 44.1 | Healthy university students | 2085 mg EPA AND 348 mg DHA | mixture of palm, olive, soy, canola, and coco butter oils | BAI, CES-D |
| Kiecolt-Glaser 2012 | 16 | 51.1 | 67.4 | Healthy, sedentary, overweight, middle-aged, and older adults | 2085 mg EPA AND 348 mg DHA OR 1043 mg EPA AND 174 mg DHA | mixture of palm, olive, soy, canola, and coco butter oils | CES-D |
| Lespérance 2011 | 8 | 46.0 | 68.5 | Patients experiencing a major depressive episode | 1050 mg EPA AND 150 mg DHA | sunflower oil | IDS, MADRS |
| Lucas 2009 | 8 | 48.8 | 100.0 | Women with moderate-to-severe psychological distress | 1050 mg EPA AND 150 mg DHA | sunflower oil | HSCL, HDRS |
| Mischoulon 2015 | 8 | 44.7 | 53.0 | Adult outpatients with major depressive disorder | 1060 mg EPA AND 274 mg DHA OR 180 mg EPA AND 900 mg DHA | soybean oil | HDRS |
| Mozaffari-Khosravi 2013 | 12 | 35.1 | 61.3 | Adult outpatients with mild-to-moderate depression | 1000 mg EPA OR 1000 mg DHA | coconut oil | HDRS |
| Rapaport 2016 | 8 | 46.1 | 58.7 | Adult outpatients with major depressive disorder | 1060 mg EPA AND 260 mg DHA OR 180 mg EPA AND 900 mg DHA | soybean oil | HDRS |
| Rondanelli 2010 | 8 | 83.9 | 100.0 | Older-adult depressed females (nursing home residents) | 1670 mg EPA AND 830 mg DHA | paraffin oil | GDS |

**EPA** eicosapentaenoic acid; **DHA** docosahexaenoic acid; **HDRS** Hamilton Rating Scale for Depression; **BDI** Beck Depression Inventory; **BAI** Beck Anxiety Inventory; **CES-D** Center for Epidemiological Studies Depression Scale; **IDS** Inventory of Depressive Symptomatology; **MADRS** Montgomery-Asberg Depression Rating Scale; **HSCL** Hopkins Symptom Checklist Depression Scale; **GDS** Geriatric Depression Scale; **HADS** Hospital Anxiety and Depression Scale

### Risk of Bias

The risk of bias is reported in Fig. 2. Some concerns, arising from the measurement of the outcome domain, were identified for five studies [48, 49, 53, 55] due to participants guessing their treatment allocation correctly, suggesting that blinding was problematic. Allocation was guessed correctly by 23% of the placebo and 24% of the fish oil group participants in Frangou et al. (2006), 28% in the placebo and 33% in the fish oil group participants in Ginty et al. (2017), 66% in the placebo and 56% in the fish oil group participants in Lucas et al. (2009), and 28.6% in the placebo and 33% in the fish oil group participants in Mozaffari-Khosravi et al. (2013). Lespérance et al. (2011) reported sufficient blinding at week 1 post-baseline; however, it is unclear whether blinding was maintained throughout the entire trial, which raises some concerns. The remaining studies were low risk of bias [50, 51, 54, 56, 57].

**Fig. 2.**
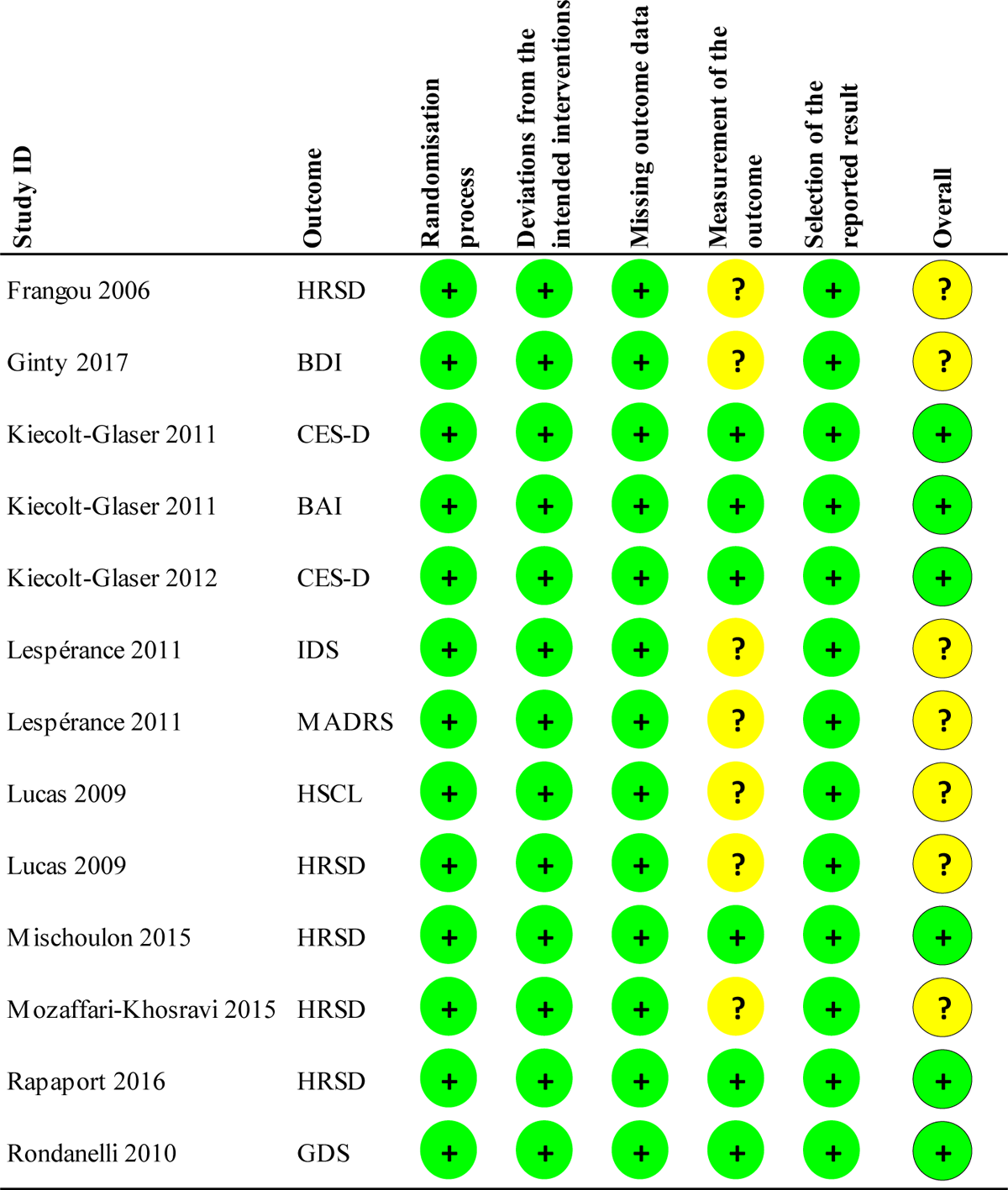
Risk of bias of included studies; 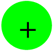 Low risk; 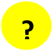 Unclear risk.

### Meta-analysis

One RCT assessed the effects of EPA and EPA-enriched, mixed EPA/DHA interventions on anxiety, reporting findings as least squares means adjusted for baseline value, visit, and gender [50]. Six RCTs assessed EPA and EPA-enriched, mixed EPA/DHA interventions on measures of depression and reported findings as mean scores at study entry and endpoint [48, 49, 52], unadjusted means at endpoint [53], endpoint means adjusted for type of antidepressant medications and baseline HDRS scores [55] and means adjusted for baseline GDS scores [57]; these were included in the same subgroup of studies reporting their findings as endpoint mean values of their respective outcome measures. Five RCTs assessed EPA-enriched, mixed EPA/DHA interventions on measures of depression and reported findings as least squares means adjusted for baseline value, visit, and gender [50], gender [51], baseline score, week of assessment, baseline antidepressants and site [52], baseline score and visit [54], and unadjusted changes from baseline to endpoint [56]. These were included in a separate subgroup of studies reporting their findings as changes from baseline values of their respective outcome measures. The non-pooled effect sizes of all the eligible studies and their respective arms included in this meta-analysis are reported in Fig. 3 1.1.6. Pooling was not performed due to different reported outcomes and different methodology in reporting outcome measures by the respective RCTs, i.e., reporting estimated means at endpoint vs. changes from baseline. Fig. 6 shows the funnel plot of the effect sizes of the RCTs, which revealed asymmetry, suggesting publication bias and heterogeneity among the trials.

**Fig. 3.**
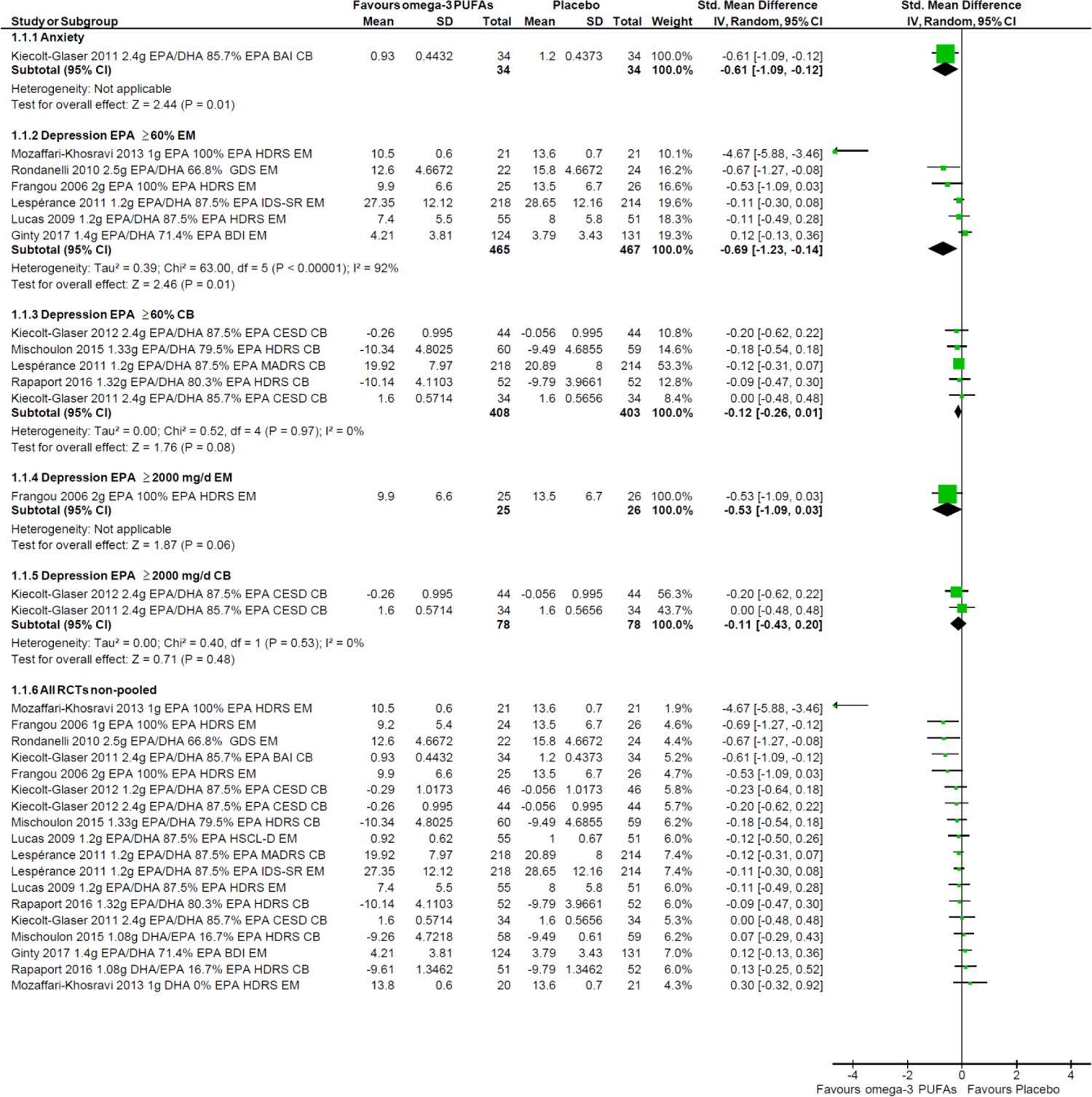
Meta-analysis of studies randomising participants to interventions comprising EPA proportions ≥60% of total EPA+DHA (1.1.2, 1.1.3) and doses ≥2000 mg/d (1.1.4, 1.1.5), and non-pooled effect studies of all the eligible trials and their respective arms (1.1.6) reporting standardised mean differences on validated anxiety and depression outcome measure scores. The trials have been categorised according to their respective reporting styles i.e., reporting endpoint mean values (EM) or change from baseline values (CB).

#### Anxiety

With only one RCT [50] assessing the efficacy against anxiety of EPA at high doses of ≥2000 mg/day and proportions ≥60% of total EPA+DHA (Fig. 3, 1.1.1) a meta-analysis could not be performed.

#### Depression

Subgroup analyses were performed comprising RCTs reporting findings as endpoint means or change from baseline on their respective outcome measures of depression, Fig. 3.

##### Effect of EPA ratio ≥60%

In the first subgroup meta-analysis for trials reporting endpoint mean values, the effects on depression of interventions with EPA proportions ≥60% of total EPA+DHA were assessed by randomising n=932 participants to a mean dose ratio of 85.5% EPA (n=465) or placebo (n=467) across six RCTs with a mean duration of 11 weeks [48, 49, 52, 53, 55, 57]. Two of these RCTs produced statistically significant results [55, 57], and the pooled estimate of effect was moderate and statistically significant (SMD: −0.69; 95% CI: −1.23, −0.14; p=0.01). This suggests that EPA-enriched interventions at proportions ≥60% of total EPA+DHA are associated with a significant reduction in depression severity, compared to placebo, by a mean of 0.69 SDs. However, between-trial heterogeneity was high; *I*^*2*^=92%, largely due to one study having a very high effect size (Fig. 3, 1.1.2). In the second subgroup meta-analysis, five RCTs with a low between-trial heterogeneity of *I*^*2*^=0% and mean duration of 10.4 weeks, that used change from baseline to assess the effects on depression of interventions comprising EPA ≥60% of total EPA+DHA, were tested by randomising n=811 participants to a mean dose ratio of 84% EPA (n=408) or placebo (n=403) [16, 50-52, 54, 56]. None of these RCTs produced statistically significant results, and the pooled estimate of effect was not statistically significant (SMD: −0.12; 95% CI: −0.26, 0.01; p=0.08) (Fig. 3, 1.1.3), which contrasts the abovementioned findings (Fig. 3, 1.1.2). However, pooling the estimates of effect of these two subgroups (1.1.2 and 1.1.3) produced statistically significant results, wherein administering interventions comprising EPA ≥60% of total EPA+DHA to a population of n=1743 participants showed a statistically significant, albeit small effect size (SMD: −0.32; 95% CI: −0.59, − 0.06; p=0.02) with considerable heterogeneity; *I*^*2*^=73.9% (Fig. 4). Of note, sensitivity analysis, comprising five out of the six RCTs in subgroup 1.1.2 [48, 49, 52, 53, 57], with Mozaffari-Khosravi et al. (2013) omitted from the analysis due to unusually large effect size, and all five RCTs in subgroup 1.1.3 [50-52, 54, 56], did not affect the above findings. Their pooled estimate of effect remained statistically significant, suggesting that the observed effect size is not substantially attributable to the outlier study by Mozaffari-Khosravi et al. (2013). However, heterogeneity for this sensitivity analysis was much reduced.

**Fig. 4.**
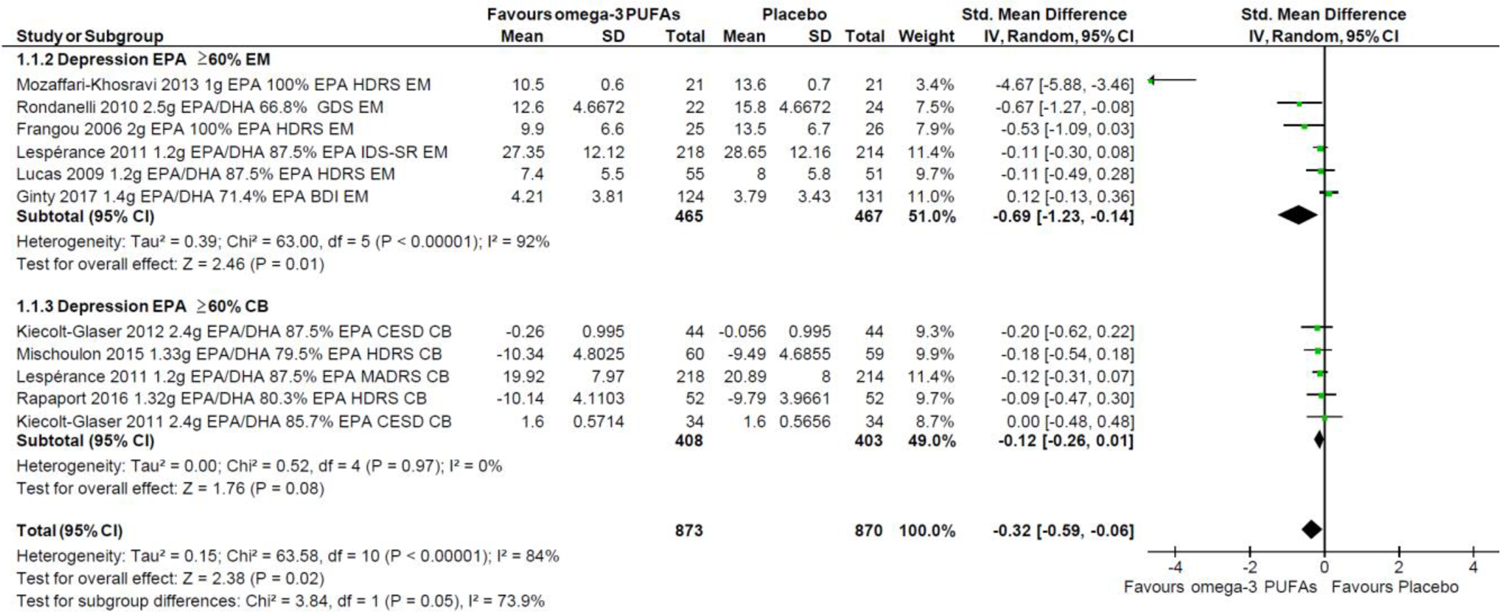
Pooled estimate of effect of subgroups 1.1.2 and 1.1.3 for randomising participants to interventions comprising ≥60% EPA ratio to DHA. The trials have been categorised according to their respective reporting styles i.e., reporting endpoint mean values (EM) or change from baseline values (CB).

##### Effect of EPA dose ≥2000 mg/day

Three RCTs assessed the efficacy against depression of EPA at high doses of ≥2000 mg/day. One of these trials reported its findings as endpoint mean values [48] (Fig. 3, 1.1.4), and two as change from baseline values [50, 51]. Therefore, a meta-analysis was only possible between the two trials using change from baseline [51, 52]. The overall effect of the two included RCTs with low between-trial heterogeneity of *I*^*2*^=0% was assessed by randomising a population of n=156 participants to a mean dose of 2085 mg/d EPA (n=78) or placebo (n=78), with a mean duration of 14 weeks. Neither of these two RCTs produced statistically significant results, and the pooled estimate of effect was not statistically significant (SMD: −0.11; 95% CI: −0.43, 0.20; p=0.48), suggesting that EPA at ≥2000 mg/day does not provide antidepressant effects (Fig. 3, 1.1.5). The single RCT testing the efficacy of EPA at high doses (≥2000 mg/day) on depression using endpoint mean values also did not show a statistically significant difference vs. placebo [49] (Fig.3, 1.1.4).

**Fig. 5.**
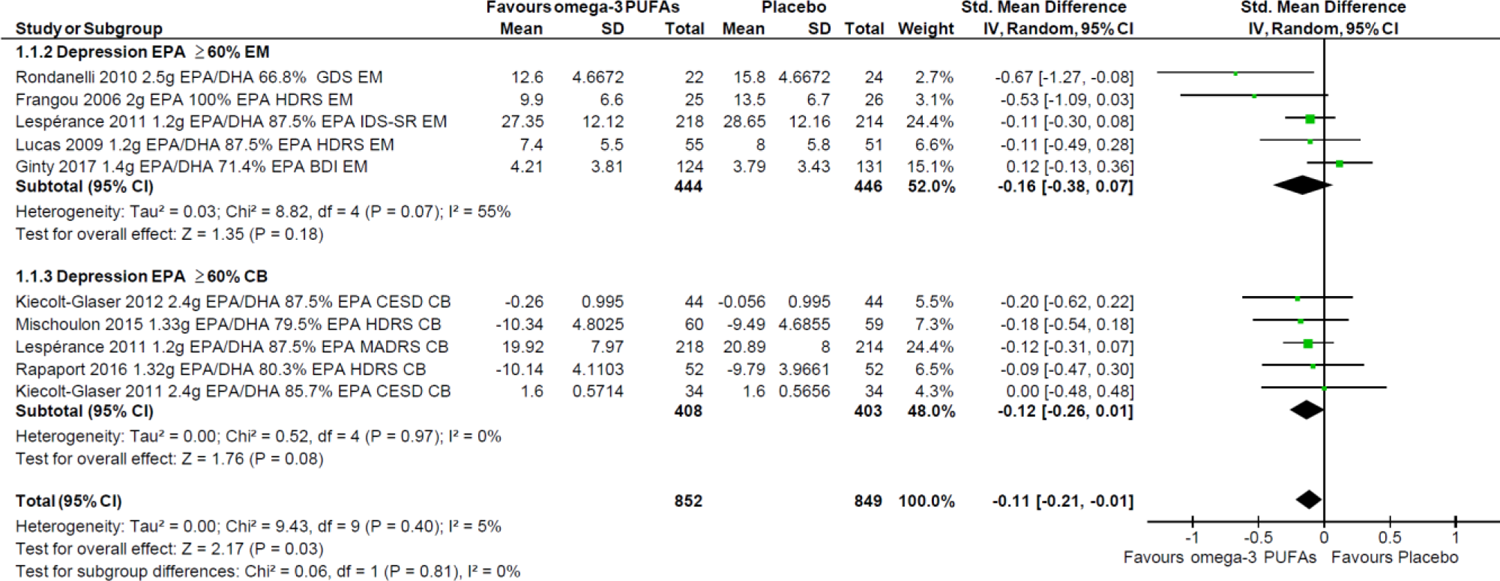
Sensitivity analysis comprising subgroups 1.1.2 and 1.1.3 for randomising participants to interventions comprising EPA proportions ≥60% of total EPA+DHA; Mozaffari-Khosravi 2013 omitted from the analysis. The trials have been categorised according to their respective reporting styles i.e., reporting endpoint mean values (EM) or change from baseline values (CB).

**Fig. 6.**
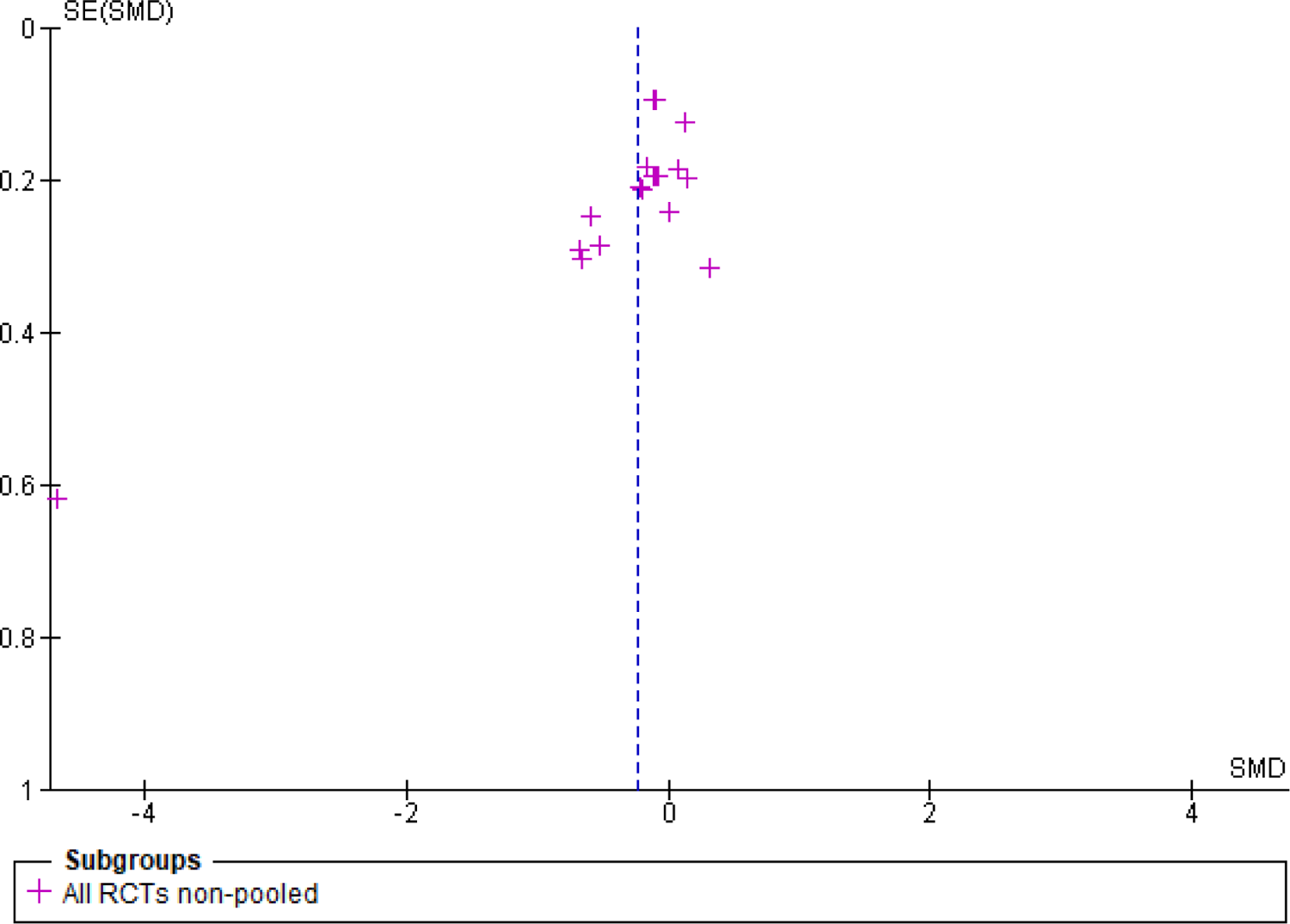
Funnel plot of the Effect Sizes of the trials included in the meta-analyses. **SE** standard error; **SMD** standardised mean differences.

## Discussion

This systematic review and meta-analysis has for the first time considered the methodological and study design limitations encountered in researching the effects of omega-3 PUFAs in anxiety and depression. The results show that EPA supplementation at proportions ≥60% of total EPA+DHA, but not at doses ≥2000 mg/day, is associated with small, but statistically significant reductions in the severity of depressive symptoms. It has also identified that no trials have investigated the antidepressant or anxiolytic effects of DPAn-3.

### Effects of EPA and DHA on depression

The antidepressant effects of EPA at proportions ≥60% of total EPA+DHA identified here are in agreement with a previous meta-analysis [20]. The statistical significance here does not necessarily imply clinical significance, and interpretation of the reported effect size should be carried out with caution, especially given the observed high heterogeneity between studies. Nonetheless, five out of six RCTs in this subgroup analysis comprised clinical populations exhibiting various degrees of severity of depression, suggesting that EPA is more efficacious against depressive symptomatology in clinical cohorts, and consequently supporting the potential clinical application of EPA.

The subgroup analysis of RCTs administering EPA doses of ≥2000 mg/day against depression did not indicate statistically significant benefit, and this contrasts with the previously reported findings by Sublette et al. (2011). The present review utilised more rigorous eligibility criteria, which were based on methodological considerations, and this reduced the number of eligible studies. Although only two studies were included in the analysis, their between-trial heterogeneity was low [50, 51], and both trials reported similar non-significant results. Both trials were of low risk of bias, thus providing quality evidence, comprised healthy adult populations, albeit of different age ranges, and administered the same doses and ratios of EPA to DHA, as well as the same depression scale (CESD). Moreover, both trials administered placebo preparations which reflected the proportions of fatty acids in the typical diet for their given populations, adding to the robustness of the findings. The EPA interventions administered in these studies were at doses of ≥2000 mg/day, but also at proportions of ≥60% of total EPA+DHA, and although this suggests that there is an upper range of therapeutic benefit, baseline severity was low for anxious and depressive symptoms in both trials, and it may be that there is less potential for improvement in healthy populations.

The confirmation of therapeutic effects with a high EPA to DHA ratio supports observations of the distinct and divergent nature of omega-3 PUFAs. The primacy of EPA in depression may be related to its anti-inflammatory effects, as unlike DHA, EPA is the precursor to prostaglandin, thromboxane, and leukotriene families of classical eicosanoids [58]. These eicosanoids are often much less biologically active than those produced from the omega-6 PUFA, arachidonic acid, and also antagonise the actions of arachidonic acid [58], which has been implicated in the pathophysiology of depression [59].

The differences in treatment efficacy between EPA and DHA against depression may also be related to their effects on neurogenesis [60]. Neurogenesis is increased by antidepressants and increasing neurogenesis has been shown to reduce anxiety and depression-related behaviours [61]. Treatment with omega-3 PUFAs also increases neurogenesis [62]; however, EPA and DHA display divergent effects on this process, whereby EPA, but not DHA, significantly increases proliferation [60]. Proliferation is induced via direct effects on the endocannabinoid signalling pathway, and there is mounting evidence supporting the role of the endocannabinoid system on the neurobiology of depression [63].

### Effects of EPA and DHA on anxiety

There was an insufficient number of RCTs investigating the efficacy of EPA and/or DHA against anxiety that met the eligibility criteria of this systematic review, and therefore a meta-analysis was not possible. However, there is literature in support of the anxiolytic properties of omega-3 PUFA interventions, and this has been reported in meta-analyses by others [19]. It is nonetheless evident, that further and more standardised research is warranted to elucidate the precise nature of the anxiolytic properties of the different omega-3 PFAs in humans, in line with recommendations for addressing methodological and study design concerns [21].

### Strengths and limitations

An important strength of this systematic review and meta-analysis is that factors that may affect the results and interpretation of the therapeutic effects of omega-3 PUFAs were considered in the eligibility criteria and search strategy syntaxes used in five key databases. For example, disease status of the participants, or presence of bioactive components in the placebo interventions. Furthermore, studies with complex interventions and multicomponent therapeutic regimes were excluded, as the potential beneficial effects of omega-3 PUFAs would be indistinguishable from those of the other components. Additionally, the authors adopted a pragmatic approach to dealing with missing data, did not contact researchers for missing values and all analyses comprised published and reported values. Where necessary, missing values were estimated using RevMan 5.4 or the web version of WebPlotDigitizer.

The strict eligibility criteria have, however, resulted in the inclusion of only a small number of RCTs comprising a heterogenous population and a small sample size, thus reflecting the need for more high-quality research in this area. In addition, baseline information on anxious and depressive outcome measure scores was not available in all the included studies, and while some RCTs reported change from baseline values, others reported their findings as endpoint mean values, making subgroup analyses necessary. This does not imply that pooled effect sizes could not be further pooled for a final estimate, but it highlights that there might be a need for standardisation in reporting findings, as highlighted elsewhere [64]. Another limitation rises from the inclusion of trials wherein blinding might have been suboptimal, which in turn raised some concerns during the assessment for risk of bias. This might be particularly problematic, considering that, due to the use of self-reporting scales to measure anxiety and/or depression, the study participants were also the outcome assessors, and experience of change in wellbeing might influence judgement in self-reported outcomes, especially where there might be strong levels of belief in either the beneficial or harmful effects of an intervention [44]. It could be argued that this is an inherent methodological limitation in the field, highlighting the importance of effective blinding, which, in turn, is a considerable challenge in the psychiatric research of omega-3 PUFAs [65]. The focus of the present systematic review and meta-analysis on placebo-controlled trials raises another methodological limitation here i.e., the putative influence of the placebo effect [66]. While it has been theorised that placebo has a robust effect on psychiatric outcomes, recent meta-analyses of RCTs demonstrated that the placebo effect in groups of anxious or depressed patients receiving placebo preparations (active controls) is small [67]. Another limitation was the lack of eligible trials addressing the efficacy of DPAn-3 against anxious and depressive symptomatology, as has been highlighted previously [8]. Finally, considering that anxiety disorders may precede the development of depression in a prodromal manner [68], the lack of sufficient eligible trials addressing the efficacy of omega-3 PUFAs against anxiety in the present systematic review and meta-analysis, but also in the majority of the therapeutic literature, is a particularly relevant limitation.

## Conclusion

In conducting this systematic review and meta-analysis many of the methodological and study design limitations encountered in research exploring the role of omega-3 PUFAs in anxiety and depression have been considered, and the results support the antidepressant effects of EPA at proportions ≥60% of total EPA+DHA in a primarily clinical population. However, EPA at doses of ≥2000 mg/day does not appear efficacious against depression severity. Further multidisciplinary research, combining psychiatry, nutrition and expertise in bioactive lipids is warranted to produce robust evidence that can be translated into clinical practice. Future studies should seek to take a more standardised approach and to address known methodological and study design limitations, in order to elucidate more fully the therapeutic potential of EPA, DHA and DPAn-3 in mental health.

## Supporting information

Search Strategy

## Data Availability

All data produced in the present work are contained in the manuscript.

https://www.crd.york.ac.uk/PROSPEROFILES/194402_STRATEGY_20201126.pdf

## Funding

This study was supported by a grant from Efamol Ltd who had no role in the study design, in the collection, analysis and interpretation of data, in the writing of the report and in the decision to submit the article for publication.

## Disclosure of interest

The authors report no conflict of interest.

## Data Availability Statement

The authors confirm that the data supporting the findings of this study are available within the article [and/or] its supplementary materials.

## Notes

### Competing Interest Statement

The authors have declared no competing interest.

### Summary of Updates

Funding Statement: This study was supported by a grant from Efamol Ltd who had no role in the study design, in the collection, analysis and interpretation of data, in the writing of the report and in the decision to submit the article for publication.

