## Supplementary material for "Effects of long-chain omega-3 polyunsaturated fatty acids on reducing anxiety and/or depression in adults; A systematic review and meta-analysis of randomised controlled trials": Search Strategy

### Search Strategies

| Source | Syntax/Search |
|---|---|
| **PubMed** | ((((((((((((((((((((((((anxiety) OR (anxi*)) OR (generalized anxiety disorder)) OR (agoraphobia)) OR (panic disorder)) OR (post-traumatic stress disorder)) OR (specific phobias)) OR (phobia)) OR (separation anxiety disorder)) OR (hypervigilance)) OR (nervousness)) OR (anxiety disorder)) OR (depression)) OR (depress*)) OR (depressive)) OR (dysphoria)) OR (dysthymia)) OR (melancholia)) OR (dysphori*)) OR (dysthym*)) OR (seasonal affective disorder*)) OR (melanchol*)) AND (((((((((((((((((((((((((eicosapentaenoic acid) OR (docosahexaenoic acids)) OR (docosapentaenoic acid)) OR (eicosapentaenoic)) OR (docosahexaenoic)) OR (docosapentaenoic)) OR (omega 3)) OR (n-3)) OR (omega-3)) OR (n3)) OR (EPA)) OR (DHA)) OR (DPA)) OR (fish oil)) OR (polyunsaturated fatty acids)) OR (PUFA)) OR (unsaturated fatty acids)) OR (highly unsaturated fatty acids)) OR (HUFA)) OR (timnodonic acid)) OR (icosapentaenoic)) OR (long chain polyunsaturated fatty acids)) OR (LC PUFAs)) OR (fish liver oils)) OR (n-3 PUFA))) AND (((((((((randomized controlled trial [pt]) OR (controlled clinical trial [pt])) OR (randomized [tiab])) OR (placebo [tiab])) OR (drug therapy [sh])) OR (randomly [tiab])) OR (trial [tiab])) OR (groups [tiab])) NOT (animals [mh] NOT humans [mh]))) Filters: Clinical Trial, Randomized Controlled Trial |
| **CINAHL** | **S1:** fish oil OR eicosapentaenoic acid OR docosahexaenoic acid OR docosapentaenoic acid OR EPA OR DHA OR DPA OR long chain omega 3 fatty acids OR long chain polyunsaturated fatty acids<br>**S2:** ( anxiety disorders or anxiety or generalized anxiety disorder ) OR anxi* OR melanchol* OR panic disorder OR ( post traumatic stress disorder or ptsd or posttraumatic stress disorder or post-traumatic stress disorder ) OR specific phobia OR separation anxiety disorder OR ( hypervigilance or nervousness ) OR ( depression or depressive disorder or depressive symptoms or major depressive disorder ) OR depress* OR dysphori* OR ( dysthymia or dysthymic disorder or persistent depressive disorder )<br>**S3:** S1 AND S2<br>Limiters - English Language; Publication Type: Randomized Controlled Trial; Age Groups: All Adult |
| **PsycInfo** | ( ( fish oil OR eicosapentaenoic acid OR docosahexaenoic acid OR docosapentaenoic acid OR EPA OR DHA OR DPA OR long chain omega 3 fatty acids OR long chain polyunsaturated fatty acids ) AND ( ( anxiety |

| | |
|---|---|
| | disorders or anxiety or generalized anxiety disorder ) OR anxi* OR melanchol* OR panic disorder OR ( post traumatic stress disorder or ptsd or posttraumatic stress disorder or post-traumatic stress disorder ) OR specific phobia OR separation anxiety disorder OR ( hypervigilance or nervousness ) OR ( depression or depressive disorder or depressive symptoms or major depressive disorder ) OR depress* OR dysphori* OR ( dysthymia or dysthymic disorder or persistent depressive disorder ) ) AND TI ( randomised control trial or randomised controlled trial or rct or randomized control trial or randomized controlled trial ) NOT TI protocol ) AND AB ( randomised control trial or randomised controlled trial or rct or randomized control trial or randomized controlled trial ) NOT TI ( injury or injuries or accident or trauma ) NOT AB ( injury or injuries or accident or trauma ) |
| **Web of Science** | **#1** (TI=(omega NEAR/3 3 NEAR/3 fatty NEAR/3 acids) OR TI=("omega-3" OR "omega 3") OR TI=(fish NEAR/1 oil*) OR TI=(eicosapentaenoic NEAR/1 acid) OR TI=(docosahexaenoic NEAR/1 acid) OR TI=(docosapentaenoic NEAR/1 acid) OR TI=(omega NEAR/4 3 NEAR/4 polyunsaturated NEAR/4 fatty NEAR/4 acids)) AND LANGUAGE: (English) Indexes=SCI-EXPANDED, SSCI, A&HCI, CPCI-S, CPCI-SSH, ESCI Timespan=All years<br>**#2** (AB=(omega NEAR/3 3 NEAR/3 fatty NEAR/3 acids) OR AB=("omega-3" OR "omega 3") OR AB=(fish NEAR/1 oil*) OR AB=(eicosapentaenoic NEAR/1 acid) OR AB=(docosahexaenoic NEAR/1 acid) OR AB=(docosapentaenoic NEAR/1 acid) OR AB=(omega NEAR/4 3 NEAR/4 polyunsaturated NEAR/4 fatty NEAR/4 acids)) AND LANGUAGE: (English) Indexes=SCI-EXPANDED, SSCI, A&HCI, CPCI-S, CPCI-SSH, ESCI Timespan=All years<br>**#3** #2 OR #1 Indexes=SCI-EXPANDED, SSCI, A&HCI, CPCI-S, CPCI-SSH, ESCI Timespan=All years<br>**#4** (TI=(anxiety) OR TI=(anxiety NEAR/1 disorder) OR TI=(depressive NEAR/1 disorder) OR TI=(depression) OR TI=(depressive) OR TI=(anxi*) OR TI=(depress*) OR TI=(mood) OR TI=(dysphori*) OR TI=(dysthymi*) OR TI=(melanchol*)) AND LANGUAGE: (English) Indexes=SCI-EXPANDED, SSCI, A&HCI, CPCI-S, CPCI-SSH, ESCI Timespan=All years<br>**#5** (AB=(anxiety) OR AB=(anxiety NEAR/1 disorder) OR AB=(depressive NEAR/1 disorder) OR AB=(depression) OR AB=(depressive) OR AB=(anxi*) OR AB=(depress*) OR AB=(mood) OR AB=(dysphori*) OR AB=(dysthymi*) |

| | |
|---|---|
| | OR AB=(melanchol*)) AND LANGUAGE: (English) Indexes=SCI-EXPANDED, SSCI, A&HCI, CPCI-S, CPCI-SSH, ESCI Timespan=All years<br>**#6** #5 OR #4 Indexes=SCI-EXPANDED, SSCI, A&HCI, CPCI-S, CPCI-SSH, ESCI Timespan=All years<br>**#7** (#3 AND #6) AND LANGUAGE: (English) AND DOCUMENT TYPES: (Article) Refined by: [excluding] DOCUMENT TYPES: ( PROCEEDINGS PAPER OR RETRACTED PUBLICATION ) AND DOCUMENT TYPES: ( ARTICLE OR EARLY ACCESS ) AND [excluding] RESEARCH AREAS: ( AGRICULTURE OR FOOD SCIENCE TECHNOLOGY OR GENERAL INTERNAL MEDICINE OR SCIENCE TECHNOLOGY OTHER TOPICS OR IMMUNOLOGY OR OBSTETRICS GYNECOLOGY OR CARDIOVASCULAR SYSTEM CARDIOLOGY OR FISHERIES OR PEDIATRICS OR CHEMISTRY OR VETERINARY SCIENCES OR ONCOLOGY OR TOXICOLOGY OR BIOTECHNOLOGY APPLIED MICROBIOLOGY OR BIOPHYSICS OR MARINE FRESHWATER BIOLOGY OR NURSING OR PUBLIC ENVIRONMENTAL OCCUPATIONAL HEALTH OR SURGERY OR ZOOLOGY OR GENETICS HEREDITY OR PHYSIOLOGY OR SPORT SCIENCES OR ENVIRONMENTAL SCIENCES ECOLOGY OR MEDICAL LABORATORY TECHNOLOGY OR RHEUMATOLOGY OR ANESTHESIOLOGY OR DEVELOPMENTAL BIOLOGY OR ENGINEERING OR MATERIALS SCIENCE OR MEDICAL INFORMATICS OR PHYSICS OR PLANT SCIENCES OR ALLERGY OR ANATOMY MORPHOLOGY OR ASTRONOMY ASTROPHYSICS OR EVOLUTIONARY BIOLOGY OR MICROBIOLOGY OR NUCLEAR SCIENCE TECHNOLOGY OR OCEANOGRAPHY OR PARASITOLOGY OR PHYSICAL GEOGRAPHY OR RADIOLOGY NUCLEAR MEDICINE MEDICAL IMAGING OR REPRODUCTIVE BIOLOGY OR THERMODYNAMICS OR TRANSPLANTATION OR UROLOGY NEPHROLOGY ) Indexes=SCI-EXPANDED, SSCI, A&HCI, CPCI-S, CPCI-SSH, ESCI Timespan=All years<br>**#8** (AB=(randomi* NEAR/2 control* NEAR/2 trial) OR TI=(randomi* NEAR/2 control* NEAR/2 trial)) AND LANGUAGE: (English) AND DOCUMENT TYPES: (Article) Indexes=SCI-EXPANDED, SSCI, A&HCI, CPCI-S, CPCI-SSH, ESCI Timespan=All years<br>**#9** #7 AND #8 Indexes=SCI-EXPANDED, SSCI, A&HCI, CPCI-S, CPCI-SSH, ESCI Timespan=All years |

| | | |
|---|---|---|
| **Cochrane Library** | **#1** | MeSH descriptor: [Fatty Acids, Omega-3] explode all trees |
| | **#2** | MeSH descriptor: [Anxiety Disorders] explode all trees |
| | **#3** | MeSH descriptor: [Depressive Disorder] explode all trees |
| | **#4** | fish oil:ti,ab,kw in Trials |
| | **#5** | MeSH descriptor: [Fish Oils] explode all trees |
| | **#6** | anxi*:ti,ab,kw in Trials |
| | **#7** | depress*:ti,ab,kw in Trials |
| | **#8** | randomi*:ti,ab,kw in Trials |
| | **#9** | mood:ti,ab,kw in Trials |
| | **#10** | omega*:ti,ab,kw in Trials |
| | **#11** | polyunsaturated:ti,ab,kw in Trials |
| | **#12** | eicosapentaenoic acid:ti,ab,kw in Trials |
| | **#13** | docosahexaenoic acid:ti,ab,kw in Trials |
| | **#14** | docosapentaenoic acid:ti,ab,kw in Trials |
| | **#15** | trial:ti,ab,kw in Trials |
| | **#16** | #1 OR #4 OR #5 OR #10 OR #11 OR #12 OR #13 OR #14 in Trials |
| | **#17** | #2 OR #3 OR #6 OR #7 OR #9 in Trials |
| | **#18** | #8 OR #15 in Trials |
| | **#19** | #16 AND #17 AND #18 in Trials |
| | **#20** | adults:ti,ab,kw in Trials |
| | **#21** | #19 AND #20 in Trials |
